# The transcriptome of CD14^+^CD163^-^HLA-DR^low^ monocytes predicts mortality in Idiopathic Pulmonary Fibrosis

**DOI:** 10.1101/2024.08.07.24311386

**Authors:** Theodoros Karampitsakos, Bochra Tourki, Minxue Jia, Carole Y. Perrot, Bogdan Visinescu, Amy Zhao, Avraham Unterman, Argyris Tzouvelekis, Debabrata Bandyopadhyay, Brenda M. Juan-Guardela, Antje Prasse, Imre Noth, Stephen Liggett, Naftali Kaminski, Panayiotis V. Benos, Jose D. Herazo-Maya

## Abstract

**Rationale:** The association between immune-cell-specific transcriptomic profiles and Idiopathic Pulmonary Fibrosis (IPF) mortality is unknown.

**Objectives:** To determine immune-cell-specific transcriptomic profiles associated with IPF mortality.

**Methods:** We profiled peripheral blood mononuclear cells (PBMC) in 18 participants [University of South Florida: IPF, COVID-19, post-COVID-19 Interstitial Lung Disease (Post-COVID-19 ILD), controls] by single-cell RNA sequencing (scRNA-seq) and identified 16 immune-cell-specific transcriptomic profiles. The Scoring Algorithm of Molecular Subphenotypes (SAMS) was used to calculate Up-scores based on these 16 gene profiles. Their association with outcomes was investigated in peripheral blood, Bronchoalveolar Lavage (BAL) and lung tissue of N=416 IPF patients from six cohorts. Findings were validated in an independent IPF, PBMC scRNA-seq dataset (N=38).

**Measurements and main results:** Cox-regression models demonstrated that 230 genes from CD14^+^CD163^-^HLA-DR^low^ circulating monocytes predicted IPF mortality [Pittsburgh (p=0.02), Chicago (p=0.003)]. PBMC proportions of CD14^+^CD163^-^HLA-DR^low^ monocytes were higher in progressive versus stable IPF (Yale, 0.13±0.05 versus 0.09±0.05, p=0.034). Receiving operating characteristic identified a 230 gene, Up-score >41.84 (Pittsburgh) predictive of mortality in Chicago (HR: 6.58, 95%CI: 2.15-20.13, p=0.001) and in pooled analysis of BAL cohorts (HR: 2.20, 95%CI: 1.44-3.37, p=0.0003). High-risk patients had decreased expression of the T-cell co-stimulatory genes *CD28*, *ICOS*, *ITK* and *LCK* (Pittsburgh and Chicago, p<0.01). 230 gene-up-scores negatively correlated with Forced Vital Capacity (FVC) in IPF lung tissues (LGRC, rho=-0.2, p=0.02). Results were replicated using a subset of 13 genes from the 230-gene signature (pooled PBMC cohorts - HR: 5.34, 95%CI: 2.83-10.06, p<0.0001).

**Conclusions:** The transcriptome of CD14^+^CD163^-^HLA-DR^low^ monocytes is associated with increased IPF mortality.

## Introduction

Idiopathic pulmonary fibrosis (IPF) represents a chronic, progressive and unpredictable lung disease(1). We have previously discovered and validated a peripheral blood 52-gene signature predictive of increased IPF mortality associated with down-regulation of the T-cell co-stimulatory genes *CD28*, *ICOS*, *ITK* and *LCK* (2, 3). Cellular deconvolution of this signature allowed the identification of classical monocyte counts as predictors of IPF mortality (2-6). Subsequent studies have corroborated the role of immune dysregulation in the pathogenesis of pulmonary fibrosis (7-9), in other fibrotic diseases and in conditions associated with increased risk of pulmonary fibrosis such as COVID-19(10). For example, a single-cell atlas of circulating immune cells in IPF demonstrated that patients with progressive IPF had an increase in monocytes and Tregs compared to controls and patients with stable disease(11). With regards to the lung tissue, the analysis of fibrotic lungs through single-cell RNA sequencing (scRNA-seq) highlighted changes in immune cell populations (12-14). While significant progress has been made regarding the identification of changes in the cell composition associated with progressive IPF, the association between immune-cell-specific gene expression profiles and Idiopathic Pulmonary Fibrosis (IPF) mortality is still unknown.

In this study, we identified sixteen immune-cell subpopulations through scRNA-seq, their cell-specific transcriptome and their mortality prediction potential in IPF patients. The transcriptome that most consistently predicted IPF mortality was the one derived from CD14^+^CD163^-^HLA-DR^low^ monocytes, cells with strong immunosuppressive capacity(15-17). A 230-gene profile and a subset of 13 genes from this profile were able to discriminate IPF patients with increased risk of mortality in PBMC and BAL and correlated with disease severity in lung tissue. Patients with a high-risk profile based on these genes also had decreased expression of the T-cell costimulatory genes *CD28*, *ICOS*, *ITK* and *LCK* suggesting an immunosuppressive effect in IPF. The 13-gene signature derived from CD14^+^CD163^-^HLA-DR^low^ monocytes can be used for risk stratification, predict treatment response and serve as the basis for future mechanistic studies aiming at modulating these monocyte-specific genes in IPF.

## Methods

### Single-cell RNA sequencing experiments

Details for the scRNA-seq experiment, analysis and quality control are provided in the **online Supplement**.

### Single-cell RNA sequencing data analysis

Data analysis revealed 16 different immune subpopulations. Subsequently, we identified the upregulated genes that were differentially expressed in each immune subpopulation compared to the other cell subpopulations (log2 Fold Change≥2.0, p<0.0001, Benjamini-Hochberg adjusted). Therefore, 16 immune-cell specific gene signatures were generated. We sought to determine which of these 16 immune-cell subpopulations are present in IPF in two independent peripheral blood scRNA-seq datasets (University of South Florida and Yale) and then we tested the prognostic performance of the transcriptome of the respective gene-signatures in two independent peripheral blood cohorts of patients with IPF (Pittsburgh and Chicago). We also tested if the gene-signature that was a significant predictor of mortality in both cohorts, retained the prognostic accuracy in BAL (Siena, Leuven and Freiburg) and lung tissue [Lung Genomics Research Consortium (LGRC)] cohorts of patients with IPF. With regards to lung tissue, we used Forced Vital Capacity (FVC) %predicted to determine the association of transcriptomics with disease severity as a surrogate of survival. Finally, we investigated if a more concise gene list, derived from the subset of genes with log2 Fold Change≥2.5 and FDR<5% for mortality in Pittsburgh, could retain the prognostic accuracy in all the aforementioned cohorts.

### Data availability

Data analyzed are available in GEO under the following GEO accession numbers: University of South Florida: GSE264196, Yale: GSE233844, Pittsburgh: GSE28221, Chicago: GSE27957, Siena, Leuven and Freiburg: GSE70867, LGRC: GSE47460.

### Ethics approval

Ethical approval for this study was given by the Institutional Review Boards / Research Integrity & Compliance of University of South Florida (Identity: Pro00032158_MODCR000003, Title: University of South Florida-Tampa General Hospital Lung Transplant Biorepository, Principal Investigator: Jose D. Herazo-Maya).

### Statistical analysis

The Scoring Algorithm of Molecular Subphenotypes (SAMS) was used to classify patients based on the level of gene expression, as previously described(3, 10). The steps of this algorithm were performed through R 2023.03.0. The detailed code is provided in the **online Supplement**. First, we calculated SAMS Up-scores based on 15 of the 16 immune-cell-specific transcriptomic profiles identified in IPF. One cell subpopulation (CD14^+^CD163^+^HLA-DR^low^ cells) was excluded from downstream analyses because it was found to be exclusively present in COVID-19 patients. We tested whether each immune-cell-specific Up-score predicted mortality in IPF using Cox regression models adjusted to Gender-Age-Physiology (GAP). Up-scores were included in the models as continuous covariates. We also implemented Receiver operating characteristic (ROC) curve for mortality to identify the optimal cut-off of the Up-score for the discrimination of patients into high and low-risk. In particular, ROC was implemented in Pittsburgh cohort and prognostication using the same threshold was validated in PBMC and BAL cohorts. In the pooled analysis, patients were pooled based on the risk stratification status they had in the respective cohort, as previously described(3). Validation was performed through Cox regression for mortality adjusted to GAP. With regards to T-cell co-stimulatory signaling pathway genes analysis, Mann-Whitney test was performed for comparisons between two groups. Spearman rank correlation was used to describe the monotonic relationship between two variables. The MedCalc v22.021 software was used for the aforementioned analysis.

## Results

### Single-cell transcriptomics reveals distinct immune cell subpopulations with differentially expressed genes in IPF

Study design is summarized in **Figure 1A**. Briefly, we performed scRNA-seq in peripheral blood mononuclear cells (PBMCs) from controls (n=4), patients with IPF (n=6), COVID-19 (n=3) and post-COVID-19 ILD (n=5) evaluated at the University of South Florida/Tampa General Hospital (n=18 samples). Patients with COVID-19 and post-COVID-19 ILD were included in this analysis since we have previously shown that genes of the 52-gene signature that was predictive of IPF mortality are also predictive of COVID-19 mortality(10). **Table 1** represents the baseline characteristics of participants in this cohort. After quality control, 92027 cells were included in scRNA-seq analysis. Sixteen (n=16) major immune cell subpopulations were identified in the University of South Florida cohort **(Figure 1B)** thus, sixteen gene-profiles were generated based on the differentially expressed genes (log2 Fold Change≥2.0, p<0.0001, Benjamini-Hochberg adjusted) with increased expression in each immune subpopulation compared to the other immune cell subpopulations. CD14^+^CD163^+^HLA-DR^low^ monocytes were detected only in patients with COVID-19 therefore, the potential of the respective gene-profile was not tested in IPF. The other fifteen immune subpopulations were present in IPF both in the University of South Florida cohort **(Figure 1B)** and in the Yale cohort **(Figure 2**, **Table 2)**. The number of differentially expressed genes with increased expression in each gene-profile derived from these 15 immune subpopulations is presented in **Table 3**.

**Figure 1.**
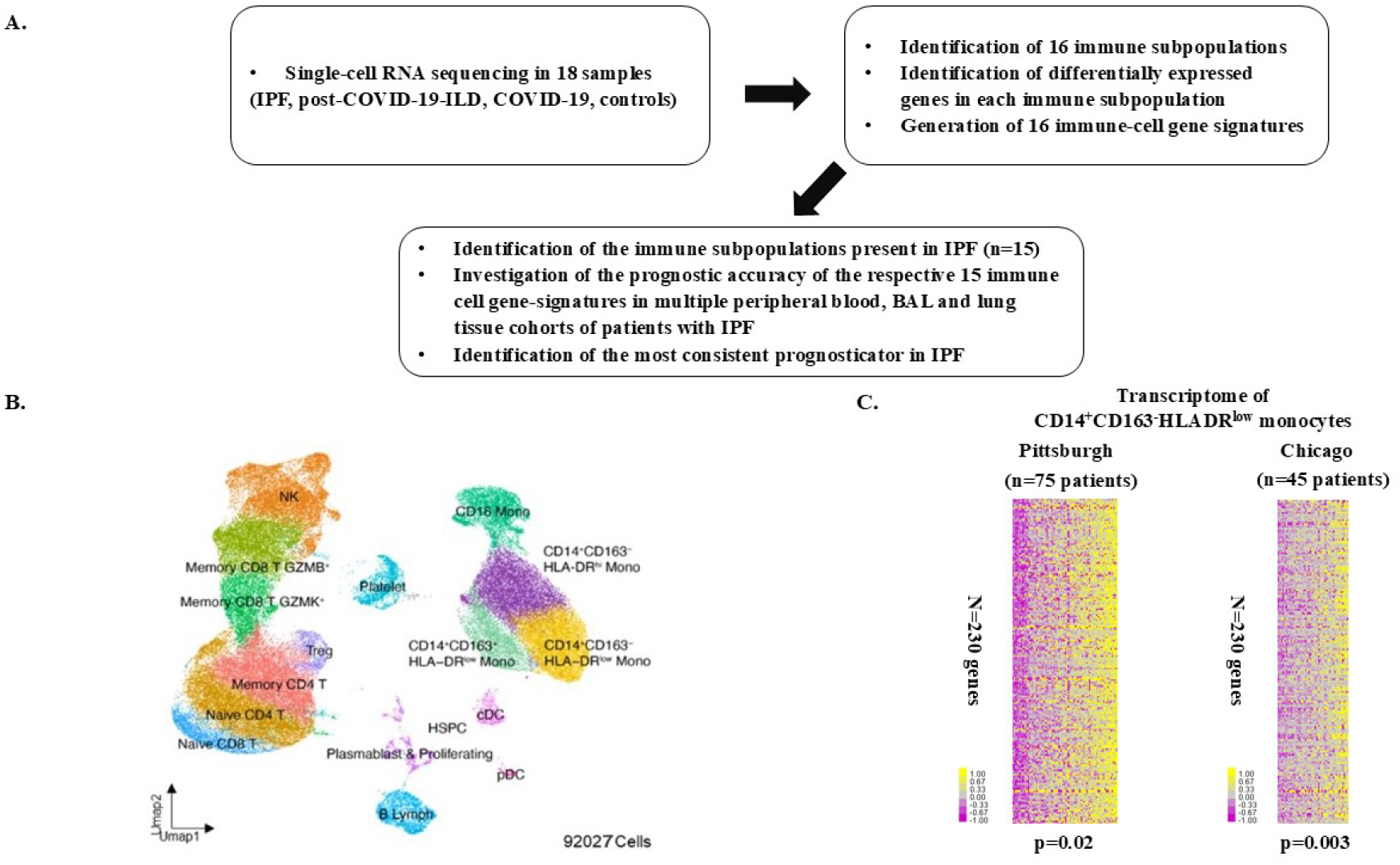
Schematic representation of the study design (Panel A). Uniform manifold approximation and projection (UMAP) embedding plots of 92027 single-cells. The cellular landscape is shown with cluster-colored annotations (Panel B). Heatmaps representing the upregulated transcriptome of CD14 CD163 HLA-DR monocytes. Every row represents a gene and every column represents a patient. Patients are sorted based on their Upscore. Color scale is shown adjacent to heat maps in log-based two scale; yellow denotes an increase over the geometric mean of samples and purple denotes a decrease. Cox regression adjusted to GAP using Upscore as a continuous covariate and mortality as endpoint was performed. The respective p values are provided underneath the heatmaps (Panel C).

**Figure 2.**
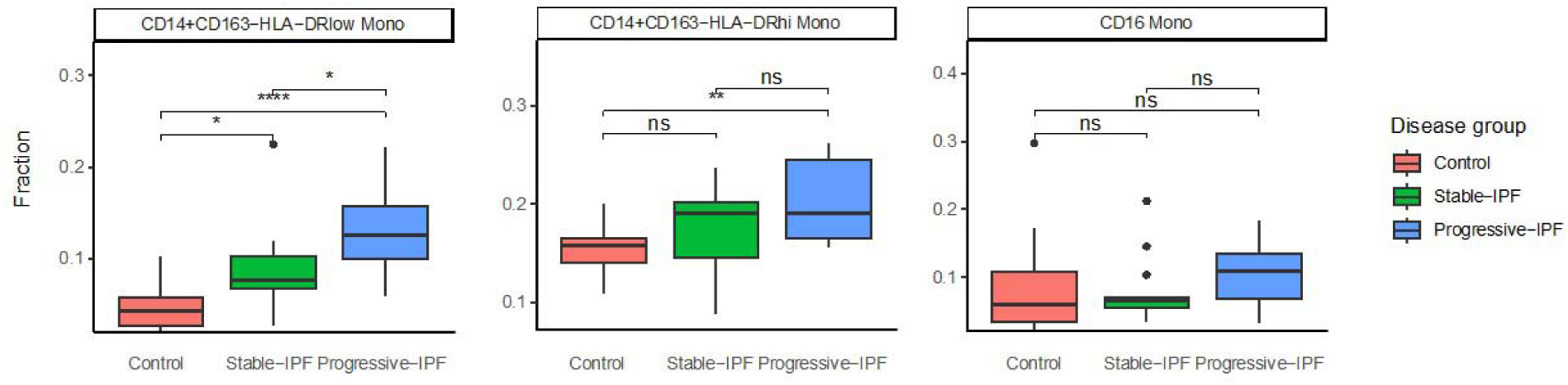
Analysis of Yale single-cell RNA sequencing dataset showed that CD14^+^CD163^-^HLA-DR^low^ monocytes was the only immune cell subpopulation with significant difference among controls, patients with stable IPF and patients with progressive IPF.

**Table 1.** Demographics and clinical data of the scRNA-seq cohort from University of South Florida.

|  | **COVID-19** | **POST COVID-19-ILD** | **IPF** | **Controls** |
| --- | --- | --- | --- | --- |
| **Number of patients** | 3 | 5 | 6 | 4 |
| **Age**<br>**(mean±SD)** | 52.0±2.6 | 61.0±12.2 | 70.0±6.4 | 42.0±2.2 |
| **Sex** |  |  |  |  |
| *Male* | 2(66.7%) | 5(83.3%) | 4(66.7%) | 2(50.0%) |
| *Female* | 1(33.3%) | 1(16.7%) | 2(33.3%) | 2(50.0%) |
| **FVC%pred**<br>**(mean±SD)** | NA | 63.8±7.2 | 61.3±14.2 | NA |
| **DLCO%pred**<br>**(mean±SD)** | NA | 44.0±2.3 | 43.8±10.1 | NA |

Abbreviations: COVID-19: Coronavirus Disease 2019, DLCO: Diffusing Capacity of the Lungs for Carbon Monoxide,
FVC: Forced Vital Capacity, ILD: Interstitial Lung Disease, IPF: Idiopathic Pulmonary Fibrosis, SD: Standard Deviation.

**Table 2.** Demographics and clinical data of the scRNA-seq cohort from Yale University.

|  | **Progressive**<br>**IPF** | **Stable**<br>**IPF** | **All**<br>**IPF** | **Controls** |
| --- | --- | --- | --- | --- |
| **Number of patients** | 12 | 13 | 25 | 13 |
| **Age**<br>**(mean±SD)** | 70.8±7.0 | 69.9±6.1 | 70.3±6.4 | 71.0±4.2 |
| **Sex** |  |  |  |  |
| *Male* | 10 (83.3%) | 10 (76.9%) | 20 (80%) | 10 (76.9%) |
| *Female* | 2 (16.7%) | 3 (23.1%) | 5 (20%) | 3 (23.1%) |
| **FVC%pred**<br>**(mean±SD)** | 69.83±11.66 | 76.85±14.62 | 73.48±13.49 | NA |
| **DLCO%pred**<br>**(mean±SD)** | 37.67±13.73 | 44.31±10.82 | 41.12±12.50 | NA |

Abbreviations: DLCO: Diffusing Capacity of the Lungs for Carbon Monoxide, FVC: Forced Vital Capacity, IPF: Idiopathic Pulmonary Fibrosis, SD: Standard Deviation.

**Table 3.**
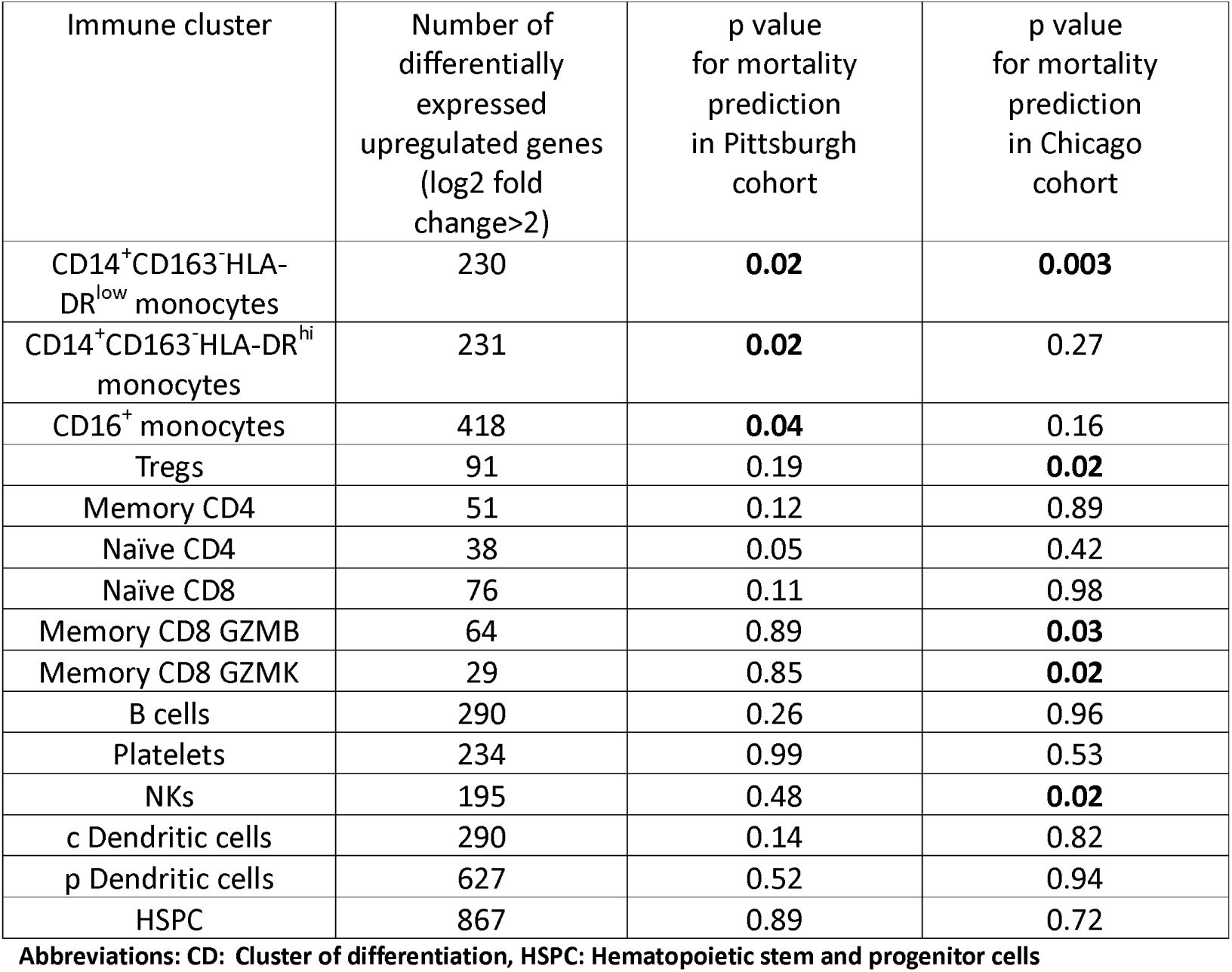
Investigation of the prognostic potential in terms of mortality prediction for the upregulated transcriptome of each immune subpopulation in peripheral blood mononuclear cells. Cox regression adjusted to GAP using Upscore as a continuous covariate was performed. The only transcriptome that consistently predicted mortality was the transcriptome of CD14 CD163 HLA-DR monocytes.

### The immune-cell-specific transcriptomic profile that consistently predicts mortality in IPF is the one derived from circulating CD14***^+^***CD163***^-^***HLA-DR***^low^***monocytes

The prognostic performance of the 15 immune-cell-specific gene expression profiles present in IPF was tested in two independent IPF cohorts: Pittsburgh (n=75 patients) and Chicago (n=45 patients), (**Table 3, Supplemental Table 2**). Demographics of these cohorts are presented in **Table 4**. Cox-regression models for mortality adjusted to GAP demonstrated that Up-scores, indicating increased expression of CD14^+^CD163^-^HLA-DR^low^ monocytes-specific genes, were independently predictive of mortality in both cohorts **(Figure 1C**, **Table 3)**. The 230 genes with increased expression in CD14^+^CD163^-^HLA-DR^low^ monocytes in IPF are listed in **Supplemental Table 1**. ROC analysis in the Pittsburgh cohort demonstrated that the optimal Up-score, cut-off threshold to discriminate patients into high or low-risk for mortality was >41.84. The prognostic performance of that threshold was tested in all PBMC and BAL cohorts. Risk stratification based on that threshold is presented in **Figure 3A** for Pittsburgh and in **Figure 3B** for Chicago, while pooled risk stratification is presented in **Figure 3C**. Cox regression adjusted to GAP demonstrated that patients in the high-risk group had increased risk for mortality compared to patients in the low-risk group both in Pittsburgh (HR: 3.00, 95%CI: 1.37-6.54, p=0.006, **Figure 3D**) and Chicago (HR: 6.58, 95%CI: 2.15-20.13, p=0.001, **Figure 3E**). Pooled analysis of Pittsburgh and Chicago cohorts also showed increased mortality risk for the high-risk group compared to the low-risk group (HR: 3.96, 95%CI: 2.12-7.37, p<0.0001, **Figure 3F).**

**Figure 3.**
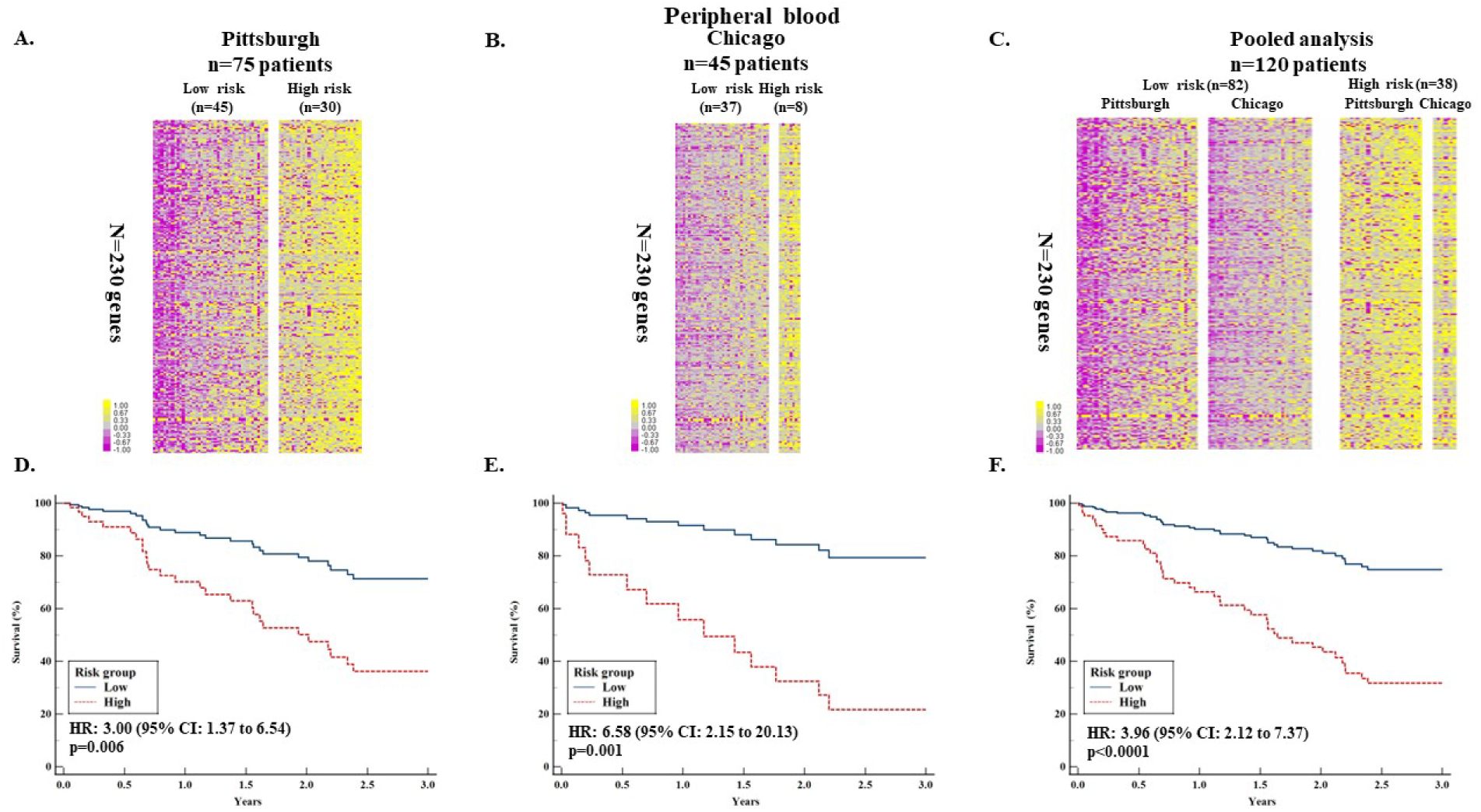
**Analysis in peripheral blood mononuclear cells:** Clustering of patients with IPF based on 230-gene risk profiles (high vs low) derived using SAMS in Pittsburgh cohort (n=75 patients). Every row represents a gene and every column represents a patient. Following SAMS calculations and receiving operating characteristic (ROC) curve, the threshold of Upscore>41.84 was used to discriminate high and low risk patients. Color scale is shown adjacent to heat maps in log-based two scale; yellow denotes an increase over the geometric mean of samples and purple denotes a decrease (Panel A). Using the same threshold, patients in Chicago cohort (n=45 patients) were split into high and low risk (Panel B). Heatmap of pooled analysis (n=120 patients), (Panel C). Cox-regression analysis adjusted to GAP showed that patients in the high-risk group had increased mortality in Pittsburgh cohort (Panel D), Chicago Cohort (Panel E) and in pooled analysis (Panel F) compared to the low-risk group.

**Table 4.** Demographics and clinical data of other cohorts included in this manuscript.

|  | **Pittsburgh** | **Chicago** | **Siena** | **Leuven** | **Freiburg** | **LGRC** |
| --- | --- | --- | --- | --- | --- | --- |
| **Number of patients** | 75 | 45 | 50 | 64 | 62 | 120 |
| **Age**<br>**(mean±SD)** | 69.0±<br>8.2 | 67.0±<br>8.1 | 68.7±<br>11.2 | 68.2±<br>8.5 | 67.4±<br>9.1 | 64.7±<br>8.3 |
| **Sex**<br>**Male**<br>**Female** | 52 (69.3%)<br>23 (30.7%) | 40 (88.9%)<br>5 (11.1%) | 40 (80.0%)<br>10 (20.0%) | 51 (79.7%)<br>13 (20.3%) | 53 (85.5%)<br>9 (14.5%) | 81 (67.5%)<br>39 (32.5%) |
| **FVC%pred**<br>**(mean±SD)** | 65.9±<br>16.8 | 61.0±<br>14.7 | 67.0±<br>23.0 | 78.0±<br>18.0 | 66.0±<br>20.0 | 66.0±<br>20.0 |
| **DLCO%pred**<br>**(mean±SD)** | 50.4±<br>19.6 | 43.3±<br>17.7 | 40.0±<br>15.0 | 45.0±<br>12 | 44.0±<br>16.0 | 48.8±<br>18.0 |

Abbreviations: DLCO: Diffusing Capacity of the Lungs for Carbon Monoxide, FVC: Forced Vital Capacity, SD: Standard Deviation.

### Single-cell RNA sequencing analysis of PBMC demonstrated that CD14 ^+^CD163^-^HLA-DR^low^ monocytes were significantly higher in patients with progressive IPF compared to patients with stable IPF and controls

We analyzed an independent PBMC scRNA-seq dataset of patients with IPF and controls from Yale University to validate our findings. Demographics of this cohort are presented in **Table 2**. Analysis of this dataset using the same markers as in the University of South Florida cohort demonstrated that CD14^+^CD163^-^HLA-DR^low^ monocytes were significantly different among all three groups: controls, stable IPF and progressive IPF (mean cell type fraction ± standard deviation - controls vs progressive IPF: 0.05±0.03 vs 0.13±0.05, p=3.6e-05, controls vs stable IPF: 0.05±0.03 vs 0.09±0.05, p=0.017, stable IPF vs progressive IPF: 0.09±0.05 vs 0.13±0.05, p=0.034).

### Patients with IPF and increased level of expression of CD14 ^+^CD163^-^HLA-DR^low^ monocyte genes have lower expression of T-cell co-stimulatory signaling pathway genes

Given that T-cell co-stimulatory signaling pathway genes predict survival in IPF(2) and that HLA-DR^low^ monocytes have been associated with features of immune paralysis in other diseases (18, 19), we sought to determine whether patients with high expression of CD14^+^CD163^-^HLA-DR^low^ monocyte genes (high-risk group) had significant differences in T-cell co-stimulatory signaling pathway genes compared to the low-risk group. Patients in the high-risk group in the Pittsburgh cohort had significantly lower expression of *CD28* [10.60 (95%CI: 10.34 to 10.82) vs 11.22 (95%CI: 11.04 to 11.34), p<0.0001]*, ICOS* [10.07 (95%CI: 9.95 to 10.28) vs 10.52 (95%CI: 10.35 to 10.61), p<0.0001]*, ITK* [11.26 (95%CI: 10.95 to 11.35) vs 11.68 (95%CI: 11.51 to 11.82), p<0.0001] and *LCK* [13.51 (95%CI: 13.32 to 13.57) vs 13.96 (95%CI: 13.88 to 14.11), p<0.0001] compared to patients in the low-risk group **(Figure 4A-D).** These findings were validated in the Chicago cohort. High-risk group patients in this cohort had significantly lower expression of *CD28* [8.19 (95%CI: 7.41 to 8.77) vs 8.95 (95%CI: 8.76 to 9.08), p=0.004]*, ICOS* [7.41 (95%CI: 6.51 to 8.09) vs 8.23 (95%CI: 7.98 to 8.39), p=0.006]*, ITK* [9.13 (95%CI: 8.59 to 9.69) vs 10.07 (95%CI: 9.80 to 10.21), p=0.002] and *LCK* [9.06 (95%CI: 8.20 to 9.56) vs 9.81 (95%CI: 9.70 to 10.02), p=0.002] compared to patients in the low-risk group **(Figure 4E-H).**

**Figure 4.**
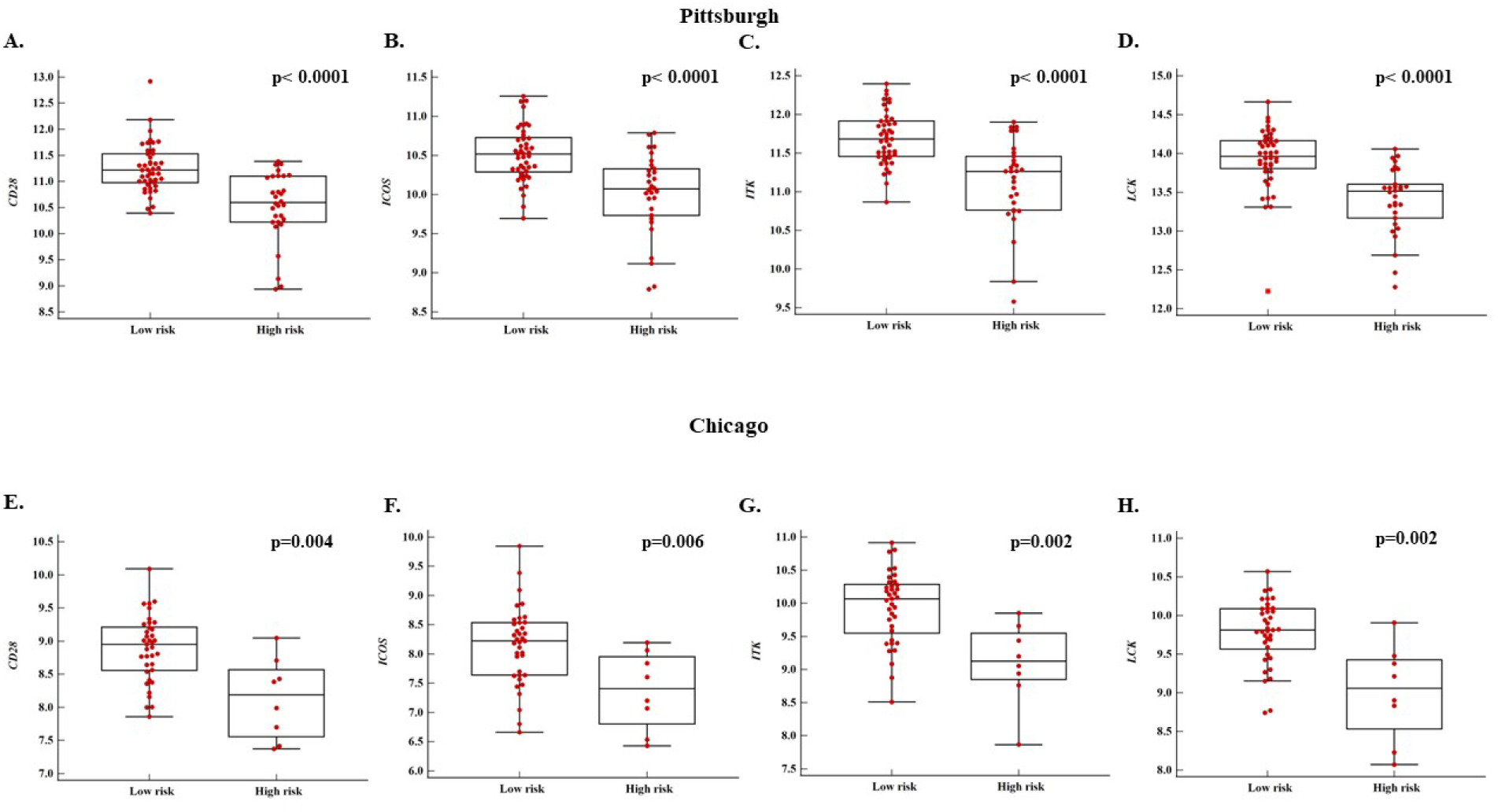
**Analysis in peripheral blood mononuclear cells:** Patients in the high-risk group had significantly lower expression of four genes belonging to the T-cell co-stimulatory signaling pathway (CD28, ICOS, ITK, LCK) compared to patients in the low-risk group both in Pittsburgh cohort (Panel A, B, C, D) and Chicago cohort (Panel E, F, G, H).

### The transcriptome of CD14 ***^+^***CD163***^-^***HLA-DR***^low^*** monocytes retains its predictive value in BAL and correlates with disease severity in lung tissue

It has been demonstrated that genes predictive of IPF mortality are expressed in circulating monocytes and can be detected in alveolar macrophages(8). Monocyte-derived alveolar macrophages (Mo-AMs) have a role in IPF progression(9, 13). Thus, we tested the prognostic performance of the immune-cell gene profile derived from *CD14*^+^*CD163*^-^*HLA-DR*^low^ *monocytes* in BAL and lung tissue to study whether alveolar macrophages, potentially derived from circulating *HLA-DR*^low^ monocytes, maintain the same mortality predictive changes in their transcriptome. Demographics of patients from these cohorts are presented in **Table 4**. Risk stratification in BAL based on the peripheral blood Up-score threshold discovered in Pittsburgh (>41.84) is presented in **Figure 5A**. Pooled analysis in three BAL datasets showed that patients with IPF in the high-risk group based on *CD1 4*^+^*CD163*^-^*HLA-DR*^low^ monocyte genes with increased expression, had increased risk of mortality compared to patients in the low-risk group (HR: 2.20, 95%CI: 1.44-3.37, p=0.0003, **Figure 5C)**. Spearman’s coefficient of rank correlation showed that Up-score in lung tissue was negatively correlated with FVC%predicted (rho=-0.2, p=0.02), (**Figure 5B**,).**D**Taken together, these findings suggest that HLA-DR^low^, monocyte-derived alveolar macrophages retain similar expression profiles as HLA-DR^low^ peripheral blood monocytes and are associated with increased mortality and disease severity in BAL and lung tissue.

**Figure 5.**
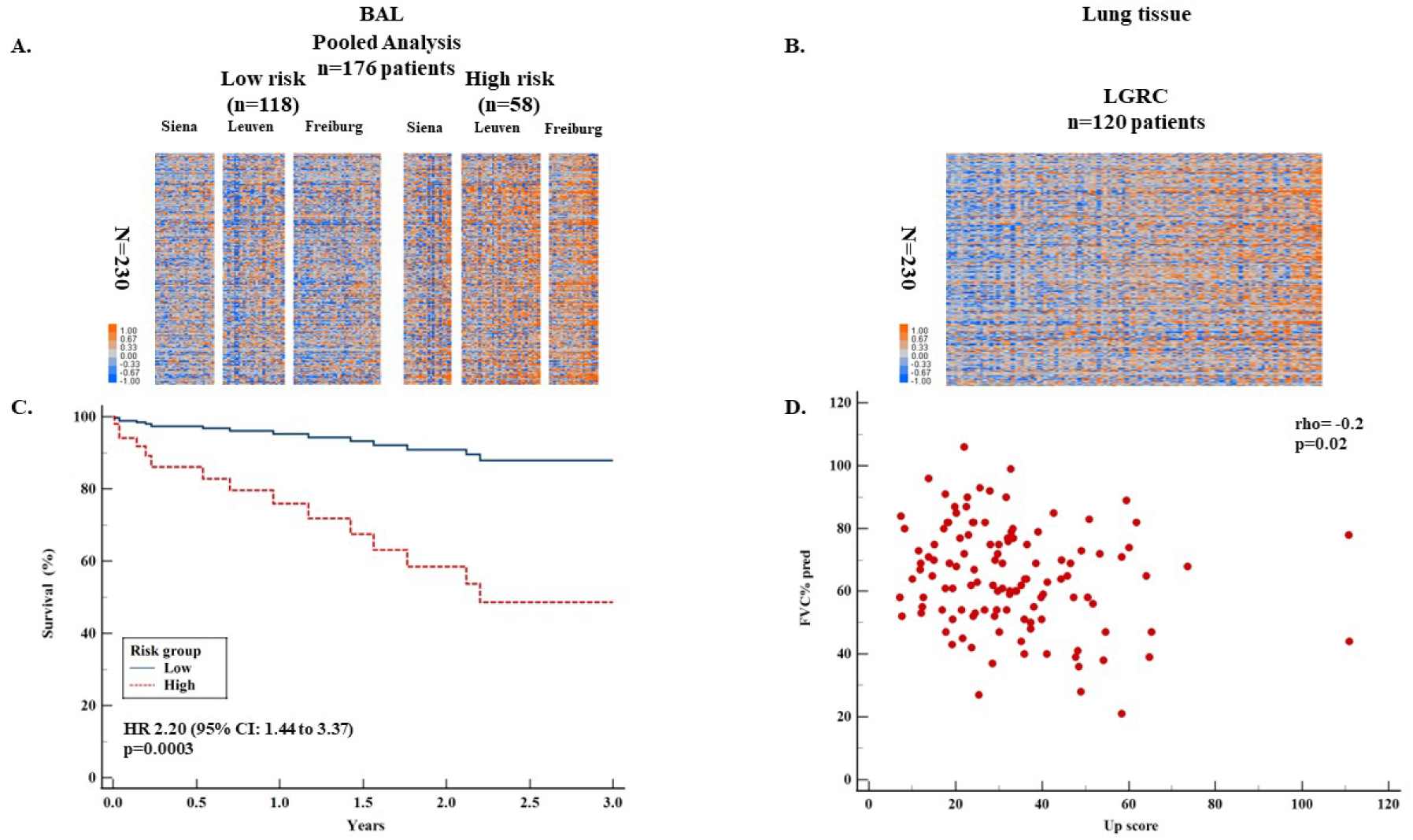
**Analysis in BAL and lung tissue:** Clustering of patients with IPF that underwent BAL (n=176 patients) based on 230-gene risk profiles (high vs low) derived using SAMS in Pittsburgh cohort. Every row represents a gene and every column represents a patient. Following SAMS calculations, the threshold of Upscore>41.84 was used to discriminate high and low risk patients. Color scale is shown adjacent to heat maps in log-based two scale; orange denotes an increase over the geometric mean of samples and blue denotes a decrease (Panel A). Accordingly, heatmap of patients with IPF that underwent surgical lung biopsy for diagnosis is presented. Patients are sorted based on the Upscore (Panel B). Cox-regression analysis adjusted to GAP demonstrated that patients in the high-risk group of the BAL cohort had increased mortality compared to the low-risk group (Panel C). Spearman’s coefficient of rank correlation shows that Upscore in lung tissue was negatively correlated with FVC%predicted (Panel D).

### A 13-gene risk profile derived from the transcriptome of CD14 ***^+^***CD163***^-^***HLA-DR***^low^***monocytes also predicts outcomes in IPF and is reproducible in peripheral blood, BAL and lung tissue

To provide a more concise and clinically applicable gene list out of the total of the 230 genes, we generated a 13-gene profile from the genes that had log2fold change>2.5 and were significantly associated with survival in the Pittsburgh cohort (FDR<5%), as previously described(2). This gene profile included *BPI, S100A12, SERINC2, ZNF697, MGST1, LIN7A, S100A9, SERPINB10, CLEC4D, CD36, S100A8, ASGR1* and *MCEMP1.* After calculating Up-scores based on the 13-gene profile, the ROC curve analysis in the Pittsburgh cohort demonstrated an Up-score threshold of >4.70 as the ideal cut-off to discriminate patients into high and low-risk of mortality. The prognostic performance of that threshold was tested in all PBMC and BAL cohorts.

Cox regression models adjusted to GAP demonstrated that patients in the high-risk group had increased risk for mortality compared to patients in the low-risk group both in the Pittsburgh (HR: 4.84, 95%CI: 2.16-10.88, p=0.0001, **Figure 6A**) and Chicago (HR: 5.41, 95%CI: 1.71-17.12, p=0.004, **Figure 6B**) PBMC, IPF cohorts. Pooled analysis in these two cohorts also showed that patients with IPF in the high-risk group exhibited worse survival compared to the low-risk group (HR: 5.34, 95%CI: 2.83-10.06, p<0.0001, **Figure 6C**). Risk stratification with the same threshold in BAL, showed that IPF patients with a high-risk profile based on these 13 genes, had increased mortality compared to the low-risk group in the BAL pooled analysis (HR: 3.72, 95%CI: 2.23-6.20, p<0.0001, **Figure 6D**). Finally, Spearman’s coefficient of rank correlation demonstrated a negative correlation between Upscore and FVC%predicted (rho=-0.2, p=0.01), (**Figure 6E**).

**Figure 6.**
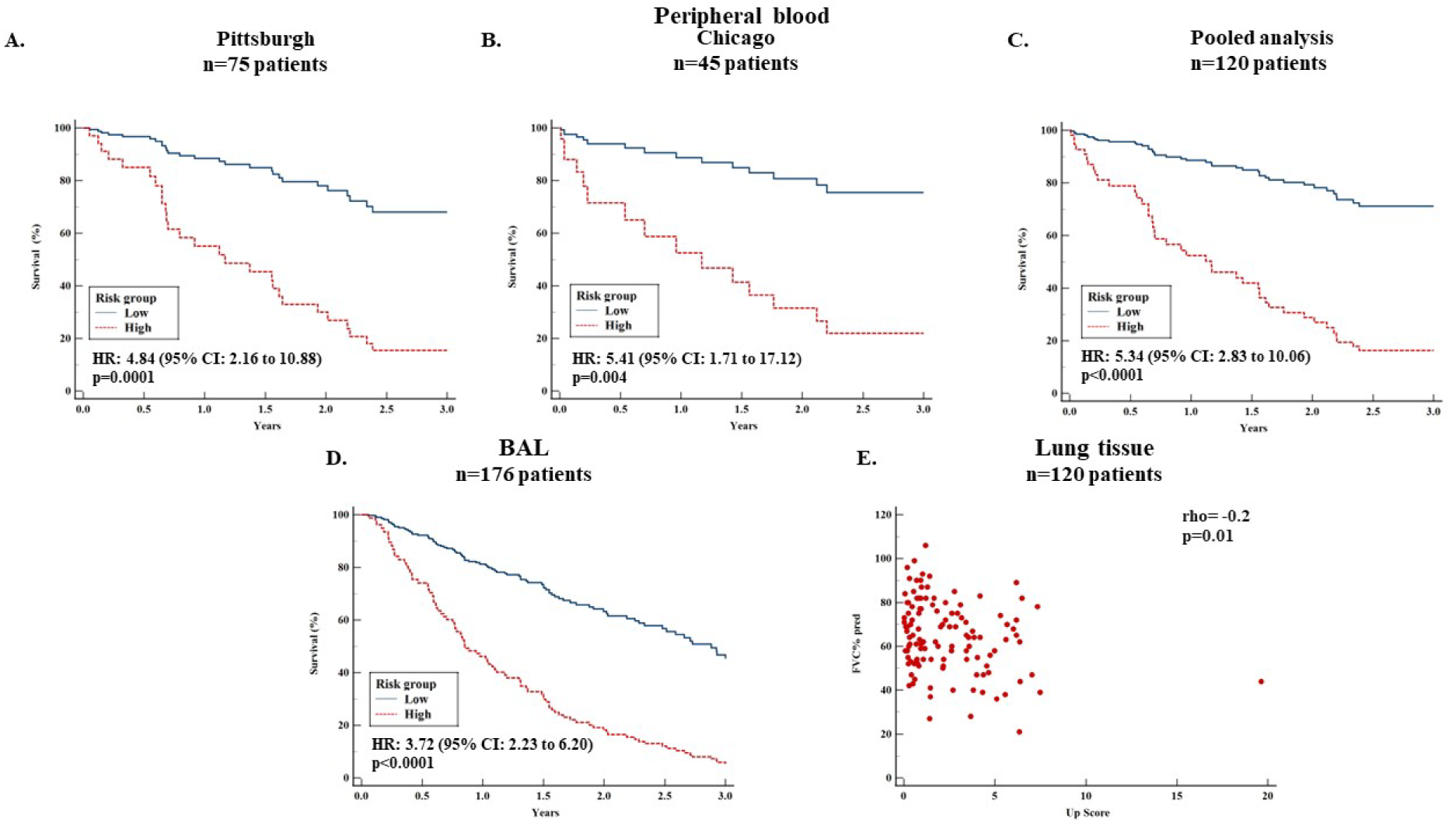
A 13-gene profile was generated from the genes that were significantly associated with survival in Pittsburgh (log2fold change>2.5 and FDR<5%) and was validated in other cohorts. This gene-profile included BPI, S100A12, SERINC2, ZNF697, MGST1, LIN7A, S100A9, SERPINB10, CLEC4D, CD36, S100A8, ASGR1 and MCEMP1 (genes were sorted based on the adjusted p value). ROC identified the value>4.70 as the ideal threshold for mortality prediction and thus was used to discriminate patients into high and low risk. Patients in the high-risk group had increased mortality risk compared to the low-risk group in Pittsburgh (Panel A), Chicago (Panel B) and in the pooled analysis (Panel C). Based on the same threshold as in the peripheral blood, patients in the high-risk group of the BAL cohort had significantly worse survival compared to the low-risk patients (Panel D). Spearman’s coefficient of rank correlation shows that Upscore of the 13-gene profile in lung tissue was negatively correlated with FVC%predicted (Panel E).

## Discussion

Our study suggests that CD14^+^CD163^-^HLA-DR^low^ monocytes are the subpopulation of monocytes driving IPF mortality and progression. Mortality and progression do not seem to be driven by the entire pool of circulating monocytes as previously described(4). This is extremely relevant because future precision medicine therapies aiming at targeting monocytes in IPF should not target the entire pool of monocytes but those associated with disease progression. The transcriptome of CD14^+^CD163^-^HLA-DR^low^ monocytes predicted poor outcomes in multiple cohorts of patients with IPF and was reproducible in peripheral blood, BAL and lung tissue. The reproducibility of this observation was shown using the entire set of differentially expressed genes with increased expression in CD14^+^CD163^-^HLA-DR^low^ monocytes (n=230 genes) and with a subset of these genes. Our findings were further corroborated by analysis of an independent PBMC scRNA-seq dataset. This analysis demonstrated that CD14^+^CD163^-^HLA-DR^low^ monocytes were significantly higher in patients with progressive IPF compared to patients with stable IPF and controls. The main attributes of our study are presented below.

We generated 16 immune-cell-specific gene profiles derived from the respective 16 immune subpopulations identified in the University of South Florida, scRNA-seq cohort. We included controls, patients with COVID-19 and post-COVID-19 due to our recent reports demonstrating similar outcome predictive profiles in peripheral blood between IPF and COVID-19(10). This approach was crucial to identify the subpopulation of classical monocytes that is present in IPF (CD14^+^CD163^-^HLA-DR^low^) and the subcluster that seems to be present in acute viral infections associated with a profibrotic risk such as COVID-19 (CD14^+^CD163^+^HLA-DR^low^). With this approach we were able to better define the differential transcriptome of each subpopulation and we generated immune-cell specific gene-signatures. Subsequently, we tested the prognostic potential of these gene-profiles in IPF through an unbiased way and particularly through a Cox-regression model in which Up-scores were treated as a continuous covariate. This led to the finding that the most consistent immune-cell-specific transcriptome predictive of IPF mortality was the one found in CD14^+^CD163^-^HLA-DR^low^ monocytes. Following that observation, we aimed to determine the optimal Up-score cut-offs to discriminate patients into high and low-risk groups and we validated the risk stratification model using this threshold in other peripheral blood and BAL cohorts.

We showed in this work that a subcluster of classical monocytes (CD14^+^CD163^-^HLA-DR^low^) is the one that consistently predicts mortality in IPF out of the entire pool of monocytes. An increase of classical monocytes in the peripheral blood of patients with IPF compared to controls has already been shown(11). Given our findings for the specific cluster of classical monocytes that drives mortality, classification of monocytes based on the approach we described might be more appropriate compared to the traditional classification of classical, intermediate and non-classical monocytes(9). Of note, one of the 13 genes of our signature, named Mast-cell expressed membrane protein-1 (*MCEMP1*), has been recently shown to be a Transforming Growth Factor-beta responsive gene that participates in monocyte chemotaxis and migration and can be identified in alveolar macrophages in IPF(8). This suggests that alveolar macrophages expressing *MCEMP1* are monocyte-derived and potentially explain the reason why the same transcriptome of CD14^+^CD163^-^ HLA-DR^low^ monocytes is able to predict mortality in the IPF and BAL datasets. Another finding supporting this concept is that C-C chemokine receptor type 2 ligands such as chemokine CC motif ligand 7 were increased in the plasma of patients with IPF and might have a role in the monocyte recruitment into the fibrotic lung(11). Other reports have also suggested that peripheral blood monocytes are precursors of the lung macrophages, which have a cardinal role in the fibrotic niche (9, 20, 21). Our observation that the transcriptome of CD14^+^CD163^-^HLA-DR^low^ monocytes retained its prognostic accuracy in BAL further supports the concept that monocyte-derived macrophages have a key role in the pathogenesis of pulmonary fibrosis(22, 23) and implies that the monocytes orchestrating this phenomenon are those with HLA-DR loss.

The effect of HLA-DR loss on the phenotype of monocytes has been studied in the setting of several diseases; yet, the precise mechanisms still need to be elucidated(24). HLA-DR is widely known as one of the three Major Histocompatibility Complex class II glycoproteins expressed on antigen-presenting cells that present peptides to T-cell receptors. The aforementioned phenomenon leads to T-cell activation. Low or no HLA-DR results into diminished capacity to present antigens to T cells (24, 25). Several lines of evidence have suggested that monocytes with HLA-DR loss, named either CD14^+^HLA-DR^lo/neg^ monocytes or monocytic Myeloid Derived Suppressor Cells, become immunosuppressive and lead to ‘’immunoparalysis’’ (24-27). This provides a plausible explanation for the reason that patients with high expression of the transcriptome of CD14^+^CD163^-^ HLA-DR^low^ monocytes had reduced expression of T-cell co-stimulatory signaling pathway genes in our study and this could also be the one the of the reasons that concomitant use of steroids and azathioprine increased mortality in IPF(28). Moreover, CD14^+^CD163^-^HLA-DR^low^ monocytes have been shown to promote expansion of Tregs and suppress Natural Killer (NK) cells(17). The expansion of Tregs and the suppression of NK cells has been reported in IPF and has been associated with disease progression (11, 20, 29-34).

Mechanistic studies aiming to identify factors that can alter the HLA-DR status demonstrated that several cytokines such as IL-1β and TGF-β could downregulate HLA-DR expression in monocytes(35, 36). Finally, expression of HLA-DR on monocytes has been suggested as a biomarker in multiple other settings such as sepsis, response to injury and response to immunotherapy (18, 37-39). This suggests that these cells and their transcriptome might have prognostic and therapeutic potential during injury and aberrant repair.

Finally, another attribute of this work is that we present for the first time a user-friendly version of SAMS through R in the online Supplement. Given that SAMS has been used in previous studies for risk stratification based on gene-expression(3, 10), we aimed to provide a quick and accurate way to use this algorithm, which may help risk-stratification in future gene expression studies as well. Computational approaches and artificial intelligence models might substantially contribute to advances in genomics and precision medicine.

Despite the reproducibility of our findings in multiple cohorts of patients, we need to acknowledge some limitations. First, we did not investigate survival differences in lung tissue due to lack of long-term survival data in this cohort. However, we used FVC% predicted as a surrogate of survival for the lung-tissue cohort. Moreover, we showed that the transcriptome retained its predictive accuracy in BAL, which also provides substantial piece of information for what happens in the lung, where the disease is. Second, we provided scRNA-seq data for the peripheral blood, but not for the lung tissue. Third, we did not study acute exacerbations of patients with IPF. Future studies could investigate if CD14^+^CD163^+^HLA-DR^low^ monocytes participate in acute exacerbations of patients with IPF, given that we identified these cells specifically in COVID-19. Finally, our study is not mechanistic in nature. Future studies aiming at demonstrating whether HLA-DR loss in monocytes associates with decreased T-cell co-stimulation and increased lung fibrosis in-vitro and in-vivo are necessary but are out of the scope of this study. Additional validation and mechanistic studies are greatly anticipated.

In conclusion, our study provided novel insights for the role of immunity in IPF. Our findings suggest that CD14^+^CD163^-^HLA-DR^low^ monocytes known to have an immunosuppressive phenotype, represent the subcluster of monocytes that drives mortality in IPF. The transcriptome of CD14^+^CD163^-^HLA-DR^low^ monocytes predicted outcomes in multiple cohorts of patients with IPF in peripheral blood, BAL and lung tissue. Thus, this transcriptome may guide risk stratification and development of new therapeutic interventions in IPF.

## Declarations

### Presentation of preliminary data

Preliminary data related to this work were presented in ATS 2024. The abstract was a recipient of ATS Abstract Scholarship.

### Data availability

Data analyzed are available in GEO under the following GEO accession numbers: University of South Florida: GSE264196, Yale: GSE233844, Pittsburgh: GSE28221, Chicago: GSE27957, Siena, Leuven and Freiburg: GSE70867, LGRC: GSE47460.

### Ethics approval

Ethical approval for this study was given by the Institutional Review Boards / Research Integrity & Compliance of University of South Florida (Identity: Pro00032158_MODCR000003, Title: University of South Florida-Tampa General Hospital Lung Transplant Biorepository, Principal Investigator: Jose D. Herazo-Maya).

### Authors approval

All authors approved this form of the manuscript.

## References

1. Raghu G, Remy-Jardin M, Richeldi L, Thomson CC, Inoue Y, Johkoh T, et al. Idiopathic Pulmonary Fibrosis (an Update) and Progressive Pulmonary Fibrosis in Adults: An Official ATS/ERS/JRS/ALAT Clinical Practice Guideline. Am J Respir Crit Care Med. 2022;205(9):e18–e47.

2. Herazo-Maya JD, Noth I, Duncan SR, Kim S, Ma SF, Tseng GC, et al. Peripheral blood mononuclear cell gene expression profiles predict poor outcome in idiopathic pulmonary fibrosis. Sci Transl Med. 2013;5(205):205ra136.

3. Herazo-Maya JD, Sun J, Molyneaux PL, Li Q, Villalba JA, Tzouvelekis A, et al. Validation of a 52-gene risk profile for outcome prediction in patients with idiopathic pulmonary fibrosis: an international, multicentre, cohort study. Lancet Respir Med. 2017;5(11):857–68.

4. Scott MKD, Quinn K, Li Q, Carroll R, Warsinske H, Vallania F, et al. Increased monocyte count as a cellular biomarker for poor outcomes in fibrotic diseases: a retrospective, multicentre cohort study. Lancet Respir Med. 2019;7(6):497–508.

5. Kreuter M, Lee JS, Tzouvelekis A, Oldham JM, Molyneaux PL, Weycker D, et al. Monocyte Count as a Prognostic Biomarker in Patients with Idiopathic Pulmonary Fibrosis. Am J Respir Crit Care Med. 2021;204(1):74–81.

6. Karampitsakos T, Torrisi S, Antoniou K, Manali E, Korbila I, Papaioannou O, et al. Increased monocyte count and red cell distribution width as prognostic biomarkers in patients with Idiopathic Pulmonary Fibrosis. Respir Res. 2021;22(1):140.

7. Shenderov K, Collins SL, Powell JD, Horton MR. Immune dysregulation as a driver of idiopathic pulmonary fibrosis. J Clin Invest. 2021;131(2).

8. Perrot CY, Karampitsakos T, Unterman A, Adams T, Marlin K, Arsenault A, et al. Mast-cell expressed membrane protein-1 is expressed in classical monocytes and alveolar macrophages in idiopathic pulmonary fibrosis and regulates cell chemotaxis, adhesion, and migration in a TGFβ-dependent manner. American Journal of Physiology-Cell Physiology. 2024;326(3):C964–C77.

9. Perrot CY, Karampitsakos T, Herazo-Maya JD. Monocytes and Macrophages: Emerging Mechanisms and Novel Therapeutic Targets in Pulmonary Fibrosis. Am J Physiol Cell Physiol. 2023.

10. Juan Guardela BM, Sun J, Zhang T, Xu B, Balnis J, Huang Y, et al. 50-gene risk profiles in peripheral blood predict COVID-19 outcomes: A retrospective, multicenter cohort study. EBioMedicine. 2021;69:103439.

11. Unterman A, Zhao AY, Neumark N, Schupp JC, Ahangari F, Cosme C, Jr., et al. Single-Cell Profiling Reveals Immune Aberrations in Progressive Idiopathic Pulmonary Fibrosis. Am J Respir Crit Care Med. 2024.

12. Reyfman PA, Walter JM, Joshi N, Anekalla KR, McQuattie-Pimentel AC, Chiu S, et al. Single-Cell Transcriptomic Analysis of Human Lung Provides Insights into the Pathobiology of Pulmonary Fibrosis. Am J Respir Crit Care Med. 2019;199(12):1517–36.

13. Morse C, Tabib T, Sembrat J, Buschur KL, Bittar HT, Valenzi E, et al. Proliferating SPP1/MERTK-expressing macrophages in idiopathic pulmonary fibrosis. Eur Respir J. 2019;54(2).

14. Juan-Guardela BM, Herazo-Maya JD. Immunity, Ciliated Epithelium, and Mortality: Are We Ready to Identify Idiopathic Pulmonary Fibrosis Endotypes With Prognostic Significance? Chest. 2022;161(6):1440–1.

15. Palojärvi A, Petäjä J, Siitonen S, Janér C, Andersson S. Low monocyte HLA-DR expression as an indicator of immunodepression in very low birth weight infants. Pediatric Research. 2013;73(1):469–75.

16. Yao R-Q, Zhao P-Y, Li Z-X, Liu Y-Y, Zheng L-Y, Duan Y, et al. Single-cell transcriptome profiling of sepsis identifies HLA-DRlowS100Ahigh monocytes with immunosuppressive function. Military Medical Research. 2023;10(1):27.

17. Lin Y, Gustafson MP, Bulur PA, Gastineau DA, Witzig TE, Dietz AB. Immunosuppressive CD14+HLA-DR(low)/- monocytes in B-cell non-Hodgkin lymphoma. Blood. 2011;117(3):872–81.

18. Mengos AE, Gastineau DA, Gustafson MP. The CD14(+)HLA-DR(lo/neg) Monocyte: An Immunosuppressive Phenotype That Restrains Responses to Cancer Immunotherapy. Front Immunol. 2019;10:1147.

19. Frazier WJ, Hall MW. Immunoparalysis and adverse outcomes from critical illness. Pediatr Clin North Am. 2008;55(3):647–68, xi.

20. Desai O, Winkler J, Minasyan M, Herzog EL. The Role of Immune and Inflammatory Cells in Idiopathic Pulmonary Fibrosis. Front Med (Lausanne). 2018;5:43.

21. Ogawa T, Shichino S, Ueha S, Matsushima K. Macrophages in lung fibrosis. International Immunology. 2021;33(12):665–71.

22. Mannes PZ, Adams TS, Farsijani S, Barnes CE, Latoche JD, Day KE, et al. Noninvasive assessment of the lung inflammation-fibrosis axis by targeted imaging of CMKLR1. Sci Adv. 2024;10(25):eadm9817.

23. Greiffo FR, Viteri-Alvarez V, Frankenberger M, Dietel D, Ortega-Gomez A, Lee JS, et al. CX3CR1-fractalkine axis drives kinetic changes of monocytes in fibrotic interstitial lung diseases. Eur Respir J. 2020;55(2).

24. Mengos AE, Gastineau DA, Gustafson MP. The CD14+HLA-DRlo/neg Monocyte: An Immunosuppressive Phenotype That Restrains Responses to Cancer Immunotherapy. Frontiers in Immunology. 2019;10.

25. Veglia F, Perego M, Gabrilovich D. Myeloid-derived suppressor cells coming of age. Nat Immunol. 2018;19(2):108–19.

26. Condamine T, Gabrilovich DI. Molecular mechanisms regulating myeloid-derived suppressor cell differentiation and function. Trends Immunol. 2011;32(1):19–25.

27. Laborde RR, Lin Y, Gustafson MP, Bulur PA, Dietz AB. Cancer Vaccines in the World of Immune Suppressive Monocytes (CD14(+)HLA-DR(lo/neg) Cells): The Gateway to Improved Responses. Front Immunol. 2014;5:147.

28. Idiopathic Pulmonary Fibrosis Clinical Research N, Raghu G, Anstrom KJ, King TE, Jr., Lasky JA, Martinez FJ. Prednisone, azathioprine, and N-acetylcysteine for pulmonary fibrosis. N Engl J Med. 2012;366(21):1968-77.

29. Huang Y, Oldham JM, Ma S-F, Unterman A, Liao S-Y, Barros AJ, et al. Blood Transcriptomics Predicts Progression of Pulmonary Fibrosis and Associated Natural Killer Cells. American Journal of Respiratory and Critical Care Medicine. 2021;204(2):197–208.

30. Moore MW, Herzog EL. Regulatory T Cells in Idiopathic Pulmonary Fibrosis: Too Much of a Good Thing? Am J Pathol. 2016;186(8):1978–81.

31. Peng X, Moore MW, Peng H, Sun H, Gan Y, Homer RJ, et al. CD4+CD25+FoxP3+ Regulatory Tregs inhibit fibrocyte recruitment and fibrosis via suppression of FGF-9 production in the TGF-β1 exposed murine lung. Front Pharmacol. 2014;5:80.

32. Cruz T, Jia M, Sembrat J, Tabib T, Agostino N, Bruno TC, et al. Reduced Proportion and Activity of Natural Killer Cells in the Lung of Patients with Idiopathic Pulmonary Fibrosis. Am J Respir Crit Care Med. 2021;204(5):608–10.

33. Cruz T, Agudelo Garcia PA, Chamucero-Millares JA, Bondonese A, Mitash N, Sembrat J, et al. End-Stage Idiopathic Pulmonary Fibrosis Lung Microenvironment Promotes Impaired NK Activity. J Immunol. 2023;211(7):1073–81.

34. Reilkoff RA, Peng H, Murray LA, Peng X, Russell T, Montgomery R, et al. Semaphorin 7a+ regulatory T cells are associated with progressive idiopathic pulmonary fibrosis and are implicated in transforming growth factor-β1-induced pulmonary fibrosis. Am J Respir Crit Care Med. 2013;187(2):180–8.

35. Gonzalez-Junca A, Driscoll KE, Pellicciotta I, Du S, Lo CH, Roy R, et al. Autocrine TGFβ Is a Survival Factor for Monocytes and Drives Immunosuppressive Lineage Commitment. Cancer Immunol Res. 2019;7(2):306–20.

36. LeibundGut-Landmann S, Waldburger JM, Krawczyk M, Otten LA, Suter T, Fontana A, et al. Mini-review: Specificity and expression of CIITA, the master regulator of MHC class II genes. Eur J Immunol. 2004;34(6):1513–25.

37. Monneret G, Lepape A, Voirin N, Bohé J, Venet F, Debard AL, et al. Persisting low monocyte human leukocyte antigen-DR expression predicts mortality in septic shock. Intensive Care Med. 2006;32(8):1175–83.

38. Ditschkowski M, Kreuzfelder E, Rebmann V, Ferencik S, Majetschak M, Schmid EN, et al. HLA-DR expression and soluble HLA-DR levels in septic patients after trauma. Ann Surg. 1999;229(2):246–54.

39. Spitzer MH, Carmi Y, Reticker-Flynn NE, Kwek SS, Madhireddy D, Martins MM, et al. Systemic Immunity Is Required for Effective Cancer Immunotherapy. Cell. 2017;168(3):487–502.e15.

