## Supplemental File 1 for "The transcriptome of CD14^+^CD163^-^HLA-DR^low^ monocytes predicts mortality in Idiopathic Pulmonary Fibrosis"

**Supplemental methods**

*Single-cell RNA sequencing experiments*

We used cryopreserved PBMCs from healthy controls and patients with IPF, COVID-19 and post-COVID-19 ILD. A single-cell suspension from PBMCs of each participant was quantified and analyzed for viability using the Cell counter 3 (Countess 3, Invitrogen). Subsequently, it was loaded into the 10X Genomics Chromium Single Cell Controller for isolation of single cells (10X Genomics). Briefly, 5000-6000 PBMC from each of the 18 samples were targeted for recovery. The single cells, reagents, and 10x Genomics gel beads were encapsulated into individual nanoliter-sized Gel beads in Emulsion (GEMs) and then reverse transcription (RT) of poly-adenylated mRNA was performed inside each droplet. Post GEM-RT Cleanup and cDNA was amplified, purified, and cDNA libraries were then prepared in bulk reactions using the Chromium Next GEM Single Cell 3ʹ Kit v3.1 Library Prep Kit. Approximately 35000 mean reads per cell were generated on the Illumina NovaSeq 1000/2000 instrument using v2.5 flow cells from sequencing. FASTQ files were generated further demultiplexing, barcode processing, alignment, and gene counting steps for analysis. Some of the aforementioned have been presented in a pre-print form(1).

**Single cell RNA-sequencing analysis and quality control**

scRNA-seq feature count matrices were constructed using Cell Ranger (v7.1.0), aligning reads to the GRCh38 2020 reference genome. Subsequent quality control and data processing were performed with the Seurat package (v4.3.0)(2)**.** Cells with less than 200 detected genes were discarded, as well as cells with more than 15% mitochondrial genes. DropletUtils (v1.14.2) identified empty droplets (3)**,** while doublets were detected using Scrublet (v0.2.3-0)(4). DecontX was used to detect ambient RNA contamination (5). Subsequently, we excluded identified empty droplets, doublets, and those with a contamination score above 0.2, alongside cells with more than 9 UMIs mapping to the hemoglobin subunit beta (HBB) gene (representing red blood cells) for further analysis. With regards to cell type annotation, we used SCTransform for data normalization and identification of variable features. This was followed by principal component analysis (PCA) and batch effect correction using Harmony (6). Graph-based clustering and UMAP embedding were generated based on Harmony embeddings. Cell types were assigned to each cluster guided by canonical marker gene expression (7-9). In the context of cell type proportions analysis, we normalized the number of cells within a given cell type by the total number of cells per subject. We used the Seurat software (v4.3.0) “FindMarkers" function to identify differentially expressed DEGs across disease groups.

**SAMS Code in R**

library(tidyverse)

library(data.table)

\# Read the 2 files

\# The First file has up to two columns (upregulated and/or downregulated genes)

\# The Second file has the raw gene expression data of patients

genes <- fread("")

raw_gene_expression <- fread("")

\# We check if columns have missing values

print("all colmuns should have no NAs")

colSums(is.na(raw_gene_expression))

\# We check that there are no duplicates

df <-raw_gene_expression %>%

group_by(Symbol) %>%

filter(n()>1)

print("genes are unique")

nrow(raw_gene_expression)==length(unique(raw_gene_expression$Symbol))

\# In case there are duplicates we remove duplicates

raw_gene_expression <- raw_gene_expression %>%

group_by(Symbol) %>%

summarise_all(first)

\# Tidy the data and mark the upregulated and downregulated

raw_gene_expression <- raw_gene_expression %>%

mutate(gene_type=if_else(Symbol%in% genes$upregulated,"upregulated",

if_else(Symbol%in%genes$downregulated,"downregulated","na"))) %>%

pivot_longer(names_to = c("Patients"),

cols =setdiff(colnames(raw_gene_expression), "Symbol"),

values_to = "Transcript")

\# Calculate Geometric mean and the Adjusted values of the geneas to the geometric mean

sams_gm <- raw_gene_expression %>%

filter(gene_type!="na") %>%

group_by(Symbol) %>%

mutate(Geom_mean=exp(mean(log(Transcript)))) %>%

mutate(Adjusted_value=Transcript-Geom_mean)

sams_adjusted <- sams_gm %>%

select(Symbol,gene_type,Patients,Adjusted_value) %>%

pivot_wider(names_from = Patients,

values_from = Adjusted_value) %>%

arrange(desc( gene_type))

fwrite(sams_adjusted, "")

\# Calculations based on percent and number of positive and negative genes

sams_score <- sams_gm %>%

group_by(Patients,gene_type) %>%

summarise(Percent_positive= sum(Adjusted_value>0)/n(),

Percent_negative= sum(Adjusted_value<0)/n(),

Nbr_positive_genes=sum(Adjusted_value>0),

Nbr_negative_genes=sum(Adjusted_value<0),

Sum_positives=sum(Adjusted_value*(Adjusted_value>0)),

Sum_negatives=sum(Adjusted_value*(Adjusted_value<0)),

.groups = 'drop'

) %>%

mutate(Up_Score=Percent_positive*Sum_positives,

Down_Score=Percent_negative*Sum_negatives) %>%

group_by(Patients) %>%

summarise(Up_Score=sum(Up_Score*(gene_type=="upregulated")),

Down_Score=sum(Down_Score*(gene_type=="downregulated")),

.groups = 'drop')

\# Risk stratification

up_score_q50 <- quantile(sams_score$Up_Score,.5)

down_score_q50 <- quantile(abs(sams_score$Down_Score),.5)

sams_risk <- sams_score %>%

mutate(Risk_up=if_else(Up_Score>up_score_q50,2,1),

Risk_down=if_else(abs(Down_Score)>down_score_q50,2,1)) %>%

mutate(Risk=Risk_up+Risk_down)

fwrite(sams_risk,"")

**Supplemental Table 1.** Genes of the 230-gene signature sorted by the level of differential expression compared to other immune subpopulations (log2fold change≥2, Benjamini-Hochberg adjusted p value<0.0001).

| Gene | avg_log2FC |
| --- | --- |
| *BPI* | 4.422772 |
| *SERPINB10* | 4.243546 |
| *HP* | 4.053879 |
| *STEAP4* | 3.981691 |
| *ADAMTS5* | 3.941835 |
| *QPCT* | 3.671006 |
| *FOLR3* | 3.600919 |
| *PADI4* | 3.506712 |
| *S100A12* | 3.467389 |
| *S100A8* | 3.447252 |
| *CLEC5A* | 3.409756 |
| *CLEC4D* | 3.380007 |
| *CMTM2* | 3.314652 |
| *SERPINB2* | 3.287744 |
| *PLA2G7* | 3.246679 |
| *PADI2* | 3.240046 |
| *CYP1B1* | 3.177962 |
| *ALDH1A1* | 3.169916 |
| *APCDD1* | 3.145837 |
| *PRLR* | 3.105551 |
| *MCEMP1* | 3.098781 |
| *GPR27* | 3.012546 |
| *KCNJ15* | 3.012155 |
| *PRICKLE1* | 3.003946 |
| *BST1* | 2.990915 |
| *RBP7* | 2.95342 |
| *S100A9* | 2.936005 |
| *TDRD9* | 2.913363 |
| *CDA* | 2.904749 |
| *LIN7A* | 2.901056 |
| *VCAN* | 2.893348 |
| *NOS1AP* | 2.87462 |
| *LPAR1* | 2.850706 |
| *S1PR3* | 2.82959 |
| *CPAMD8* | 2.828598 |
| *SPOCK3* | 2.826754 |
| *ZNF697* | 2.812669 |
| *LOXHD1* | 2.80592 |
| *MGST1* | 2.786377 |
| *DGAT2* | 2.773469 |
| *VNN3* | 2.765982 |
| *NLRP12* | 2.762679 |
| *RPH3A* | 2.752472 |
| *SPOCK1* | 2.748776 |
| *VNN2* | 2.727683 |
| *TMEM176A* | 2.727079 |
| *CYP27A1* | 2.717179 |
| *DYSF* | 2.715801 |
| *CSTA* | 2.711955 |
| *TNFAIP6* | 2.706276 |
| *NRG1* | 2.688184 |
| *TRPV4* | 2.677292 |
| *AQP9* | 2.663308 |
| *ASGR1* | 2.643851 |
| *RETN* | 2.639375 |
| *TNFRSF10C* | 2.627945 |
| *G0S2* | 2.625677 |
| *PROK2* | 2.622272 |
| *MNDA* | 2.58865 |
| *FXYD6* | 2.583837 |
| *CD14* | 2.576122 |
| *ALDH2* | 2.573285 |
| *SERINC2* | 2.571798 |
| *CD36* | 2.560781 |
| *CSF3R* | 2.555156 |
| *SEMA3C* | 2.547932 |
| *HOMER3* | 2.534212 |
| *CKAP4* | 2.525233 |
| *LGALS12* | 2.517227 |
| *CD93* | 2.513834 |
| *APBB2* | 2.506929 |
| *RGS2* | 2.496913 |
| *TGFBI* | 2.496581 |
| *MTMR11* | 2.493391 |
| *TMEM176B* | 2.489253 |
| *TMEM144* | 2.481797 |
| *GCA* | 2.480373 |
| *PYGL* | 2.475911 |
| *DSC2* | 2.469858 |
| *CLTCL1* | 2.469092 |
| *VEGFA* | 2.4653 |
| *TREM1* | 2.458981 |
| *CLMN* | 2.449941 |
| *F5* | 2.433945 |
| *BASP1* | 2.43306 |
| *NFE2* | 2.432022 |
| *CHST13* | 2.42043 |
| *FPR2* | 2.418504 |
| *CREB5* | 2.418436 |
| *FCN1* | 2.417668 |
| *SLC24A4* | 2.414685 |
| *FCAR* | 2.408402 |
| *CR1* | 2.406892 |
| *HK2* | 2.406726 |
| *SLITRK4* | 2.402962 |
| *CRISPLD2* | 2.398451 |
| *PTX3* | 2.386659 |
| *DACH1* | 2.385694 |
| *PLAUR* | 2.38448 |
| *STK32B* | 2.383921 |
| *CLEC4E* | 2.381709 |
| *PRRG4* | 2.371536 |
| *CACNA2D3* | 2.371204 |
| *FPR1* | 2.362895 |
| *LRG1* | 2.358166 |
| *PGD* | 2.340107 |
| *SLC26A8* | 2.339431 |
| *ANKRD22* | 2.337671 |
| *SIRPA* | 2.337297 |
| *CCDC149* | 2.33245 |
| *SMARCD3* | 2.328224 |
| *IL31RA* | 2.322897 |
| *ANPEP* | 2.320572 |
| *ZNF467* | 2.317179 |
| *JAG1* | 2.314646 |
| *PLBD1* | 2.306728 |
| *TMTC2* | 2.305377 |
| *CDC42EP1* | 2.29937 |
| *OSCAR* | 2.298741 |
| *RNF150* | 2.298452 |
| *OAF* | 2.297989 |
| *SMPDL3A* | 2.294219 |
| *CLEC4G* | 2.293777 |
| *F2RL1* | 2.29201 |
| *P4HA2* | 2.288629 |
| *IL1RN* | 2.282232 |
| *CLEC12A* | 2.2731 |
| *CYBB* | 2.272226 |
| *NCF2* | 2.266633 |
| *KIF13A* | 2.266326 |
| *PTGS2* | 2.258355 |
| *LRRK2* | 2.255399 |
| *ZC3H12C* | 2.250557 |
| *RAB3D* | 2.245194 |
| *TSHZ3* | 2.244426 |
| *KCNE3* | 2.229958 |
| *DOK3* | 2.229628 |
| *HLX* | 2.227015 |
| *SIGLEC15* | 2.22112 |
| *SCARF1* | 2.219435 |
| *MS4A6A* | 2.218671 |
| *SLC22A4* | 2.215183 |
| *ANKRD50* | 2.213632 |
| *WDR49* | 2.203887 |
| *SERPINI2* | 2.203833 |
| *CMTM4* | 2.203438 |
| *KBTBD11* | 2.202549 |
| *TNFSF13B* | 2.200559 |
| *CD300LB* | 2.195963 |
| *DGKG* | 2.195905 |
| *PPM1H* | 2.19527 |
| *S100P* | 2.195018 |
| *SLC22A16* | 2.188618 |
| *TRIM7* | 2.187692 |
| *CLEC4A* | 2.183326 |
| *CSF2RB* | 2.181354 |
| *HNMT* | 2.178784 |
| *ST3GAL6* | 2.17123 |
| *SLC46A2* | 2.167607 |
| *ENTPD1* | 2.16753 |
| *FCGR1B* | 2.16417 |
| *TNFSF13* | 2.157732 |
| *NDST1* | 2.157117 |
| *SIRPB1* | 2.154243 |
| *RBM47* | 2.153958 |
| *COLEC12* | 2.152489 |
| *KREMEN1* | 2.151792 |
| *SCPEP1* | 2.151226 |
| *CD33* | 2.150112 |
| *RNASE4* | 2.147095 |
| *PRAM1* | 2.143013 |
| *SIRPB2* | 2.141181 |
| *PTAFR* | 2.139879 |
| *RFX2* | 2.138331 |
| *HAL* | 2.136044 |
| *ASGR2* | 2.133129 |
| *CD1D* | 2.126497 |
| *SLC2A9* | 2.120142 |
| *SMPDL3B* | 2.117355 |
| *GK* | 2.114632 |
| *GPR162* | 2.111181 |
| *NEBL* | 2.110573 |
| *DUSP3* | 2.107764 |
| *PLCD3* | 2.105779 |
| *OLIG1* | 2.102802 |
| *MBOAT2* | 2.101134 |
| *SGK1* | 2.097138 |
| *ASRGL1* | 2.092529 |
| *TLR8* | 2.092458 |
| *WDFY3* | 2.09091 |
| *STX3* | 2.089266 |
| *NCF4* | 2.08872 |
| *RUSC2* | 2.087975 |
| *VENTX* | 2.084786 |
| *NOL3* | 2.083292 |
| *DMXL2* | 2.078998 |
| *CLEC1A* | 2.070817 |
| *MPEG1* | 2.067294 |
| *RIN2* | 2.065568 |
| *NFAM1* | 2.065546 |
| *RNASE6* | 2.064469 |
| *PSRC1* | 2.062838 |
| *KLF4* | 2.05773 |
| *FBN2* | 2.057629 |
| *VNN1* | 2.056431 |
| *TIMP2* | 2.053948 |
| *GBGT1* | 2.050828 |
| *CD300LF* | 2.047597 |
| *CTSS* | 2.046283 |
| *UBE2D1* | 2.046111 |
| *AZU1* | 2.043661 |
| *NSUN7* | 2.043473 |
| *RAB20* | 2.04045 |
| *LILRA5* | 2.040306 |
| *ARHGEF10L* | 2.03701 |
| *LILRB3* | 2.029726 |
| *NOD2* | 2.028194 |
| *APLP2* | 2.026206 |
| *ARHGEF11* | 2.025548 |
| *IMPA2* | 2.024951 |
| *DNER* | 2.022354 |
| *UBTD1* | 2.02161 |
| *CCR1* | 2.021003 |
| *CXCL2* | 2.019902 |
| *NAIP* | 2.019053 |
| *TLR2* | 2.016259 |
| *RHOU* | 2.00813 |
| *HK3* | 2.007372 |
| *MICAL2* | 2.005385 |
| *SLC6A12* | 2.00103 |

**Supplemental Table 2.** Further results of the Cox regression model adjusted to GAP using Up score as a continuous covariate. In each case, analysis included the Upscore of the respective transcriptome as a continuous covariate and GAP score.

| Immune cluster | HR (95%CI),  p value  of the transcriptome  for mortality prediction  in Pittsburgh  cohort | HR (95%CI),  p value  of GAP  for mortality prediction  in Pittsburgh  cohort | HR (95%CI),  p value  of the transcriptome  for mortality prediction  in Chicago  cohort | HR (95%CI),  p value  of GAP  for mortality prediction  in Chicago  cohort |
| --- | --- | --- | --- | --- |
| CD14^+^CD163^-^HLADR^low^ monocytes | 1.01 (1.00-1.02),   **p=0.02** | 1.63 (1.28-2.08),  **p=0.0001** | 1.02 (1.01-1.04)  **p=0.003** | 1.75 (1.23-2.49)  **p=0.002** |
| CD14^+^CD163^-^HLADR^hi^ monocytes | 1.01 (1.00-1.03)  **p=0.02** | 1.64 (1.29-2.08)  **p=0.0001** | 1.02 (0.98-1.05)  p=0.27 | 1.70 (1.22-2.38)  **p=0.002** |
| CD16^+^ monocytes | 1.01 (1.00-1.02)  **p=0.04** | 1.68 (1.31-2.15)  **p<0.0001** | 1.02 (0.99-1.06)  p=0.16 | 1.71 (1.23-2.39)  **p=0.002** |
| Tregs | 0.98 (0.95-1.01)  p=0.19 | 1.60 (1.25-2.03)  **p=0.0001** | 0.78 (0.63-0.96)  **p=0.02** | 1.71 (1.23-2.38)  **p=0.002** |
| Memory CD4 | 0.98 (0.94-1.00)  p=0.12 | 1.60 (1.26-2.03)  **p=0.0001** | 1.01 (0.89-1.15)   p=0.89 | 1.68 (1.20-2.35)  **p=0.002** |
| Naïve CD4 | 0.94 (0.89-1.00)  p=0.05 | 1.56 (1.15-2.12)  **p=0.004** | 0.92 (0.76-1.13)  p=0.42 | 1.71 (1.17-2.50)  **p=0.006** |
| Naïve CD8 | 0.97 (0.93-1.01)  0.11 | 1.59 (1.25-2.02)  **p=0.0001** | 1.00 (0.82-1.22)  p=0.98 | 1.67 (1.21-2.32)  **p=0.002** |
| Memory CD8 GZMB | 1.00 (0.98-1.02)  p=0.89 | 1.63 (1.29-2.07)  **p=0.0001** | 0.87 (0.77-0.98)  **p=0.03** | 1.89 (1.37-2.61)  **p=0.0001** |
| Memory CD8 GZMK | 0.94 (0.51-1.74)  p=0.85 | 1.64 (1.30-2.08)  **p<0.0001** | 0.59 (0.38-0.91)  **0.02** | 2.02 (1.40-2.92)  **p=0.0002** |
| B cells | 0.99 (0.99-1.00)  p=0.26 | 1.66 (1.31-2.10)  **p<0.0001** | 1.00 (0.98-1.03)  p=0.96 | 1.67 (1.21-2.31)  **p=0.002** |
| Platelets | 1.00 (1.00-1.00)  p=0.99 | 1.64 (1.30-2.07)  **p<0.0001** | 1.00 (0.99-1.01)  p=0.53 | 1.66 (1.19-2.30)  **p=0.003** |
| NKs | 1.00 (0.99-1.00)  p=0.48 | 1.67 (1.31-2.12)  **p<0.0001** | 0.27 (0.09-0.81)  **p=0.02** | 1.94 (1.36-2.76)  **p=0.0002** |
| c Dendritic cells | 1.01 (1.00-1.03)  p=0.14 | 1.67 (1.30—2.13)  **p<0.0001** | 1.12 (0.43-2.92)  p=0.82 | 1.66 (1.20-2.30)  **p=0.002** |
| p Dendritic cells | 1.00 (0.99-1.01)  0.52 | 1.65 (1.30-2.10)  **p<0.0001** | 1.00 (0.96-1.03)  p=0.94 | 1.67 (1.20-2.32)  **p=0.002** |
| HSPC | 1.00 (1.00-1.00)  p=0.89 | 1.64 (1.30-2.08)  **p<0.0001** | 1.00 (0.99-1.02)  p=0.72 | 1.64 (1.17-2.30)  **p=0.004** |

**References**

1.	Tourki B, Jia M, Karampitsakos T, Vera IM, Arsenault A, Marlin K, et al. A 50-gene high-risk profile predictive of COVID-19 and Idiopathic Pulmonary Fibrosis mortality originates from a genomic imbalance in monocyte and T-cell subsets that reverses in survivors with post-COVID-19 Interstitial Lung Disease. bioRxiv. 2023:2023.10.22.563156.

2.	Hao Y, Hao S, Andersen-Nissen E, Mauck WM, 3rd, Zheng S, Butler A, et al. Integrated analysis of multimodal single-cell data. Cell. 2021;184(13):3573-87.e29.

3.	Lun ATL, Riesenfeld S, Andrews T, Dao TP, Gomes T, Marioni JC, et al. EmptyDrops: distinguishing cells from empty droplets in droplet-based single-cell RNA sequencing data. Genome Biology. 2019;20(1):63.

4.	Wolock SL, Lopez R, Klein AM. Scrublet: Computational Identification of Cell Doublets in Single-Cell Transcriptomic Data. Cell Syst. 2019;8(4):281-91.e9.

5.	Yang S, Corbett SE, Koga Y, Wang Z, Johnson WE, Yajima M, et al. Decontamination of ambient RNA in single-cell RNA-seq with DecontX. Genome Biology. 2020;21(1):57.

6.	Korsunsky I, Millard N, Fan J, Slowikowski K, Zhang F, Wei K, et al. Fast, sensitive and accurate integration of single-cell data with Harmony. Nat Methods. 2019;16(12):1289-96.

7.	Szabo PA, Levitin HM, Miron M, Snyder ME, Senda T, Yuan J, et al. Single-cell transcriptomics of human T cells reveals tissue and activation signatures in health and disease. Nature Communications. 2019;10(1):4706.

8.	Unterman A, Sumida TS, Nouri N, Yan X, Zhao AY, Gasque V, et al. Single-cell multi-omics reveals dyssynchrony of the innate and adaptive immune system in progressive COVID-19. Nat Commun. 2022;13(1):440.

9.	Wendisch D, Dietrich O, Mari T, von Stillfried S, Ibarra IL, Mittermaier M, et al. SARS-CoV-2 infection triggers profibrotic macrophage responses and lung fibrosis. Cell. 2021;184(26):6243-61.e27.
